# Virtual dynamic contrast enhanced breast MRI using 2D U-Net Architectures

**DOI:** 10.1101/2024.08.07.24311608

**Authors:** Hannes Schreiter, Jessica Eberle, Lorenz A. Kapsner, Dominique Hadler, Sabine Ohlmeyer, Ramona Erber, Julius Emons, Frederik B. Laun, Michael Uder, Evelyn Wenkel, Sebastian Bickelhaupt, Andrzej Liebert

## Abstract

Breast Magnetic Resonance Imaging (MRI) examinations routinely include contrast-agent based dynamic contrast-enhanced (DCE) acquisitions. Expanding the accessibility and personalization of breast MRI might be supported amongst others by advancing non-contrast-enhanced MRI, such as virtual dynamic contrast-enhanced techniques (vDCE) utilizing neural networks. This IRB-approved retrospective study includes n=540 breast MRI examinations acquired on a single 3T MRI scanner. Two 2D U-Net architectures were trained using non-contrast-enhanced MRI acquisitions including T1w, T2w and multi-b-value diffusion weighted imaging acquisitions as inputs and either a single (SCO-Net) or multiple (MCO-Net) time points of a DCE series as ground truth. The neural networks predicted a vDCE series corresponding to five consecutive DCE time points. Across all time points, no significant differences in structural similarity index (SSIM) could be found between the SCO-Net and MCO-Net, both achieving a mean SSIM of 0.86. For peak-signal-to-noise-ratio and normalized root-mean-square error, significantly better results could be observed for the MCO-Net reaching scores of 24.42dB and 0.087 respectively. Comparison of manual segmentations of findings on DCE and vDCE images reached a DICE score of 0.61 and an intersection over union (IoU) of 0.47 without significant differences between SCO-Net and MCO-Net. These findings suggest a technical feasibility of generating vDCE image series from unenhanced input acquisitions using neural networks. However, the analysis does not allow drawing any conclusion on the clinical assessment of lesion specific curve kinetics, which need to be assessed prior determining on the feasibility of deriving diagnostically meaningful enhancement characteristics in individual lesions.

## 1 Introduction

Dynamic contrast-enhanced (DCE) Magnetic Resonance Imaging (MRI), a core element of multiparametric breast MRI examinations, acquires several T1-weighted (T1w) images before and after gadolinium-based contrast agent (GBCA) administration. This acquisition commonly employs intervals of 60-90 seconds in between the acquisitions, enabling assessment of enhancement characteristics in tissue and suspicious lesions [1]. Despite its diagnostic accuracy, DCE MRI has certain drawbacks when considered for specific diagnostic applications, especially in the context of breast cancer screening in healthy persons: GBCA administration is associated to direct and indirect costs [2], multi-time point DCE breast examinations commonly occupy significant scanner time [3,4] and despite a contextual comparatively high safety profile, GBCA administration have been reported to be associated with certain side effects like allergic reactions and the “symptoms associated to gadolinium exposure” (SAGE) complex [5]. Additionally, the use of GBCAs contributes to environmental gadolinium pollution with GBCA contamination affecting surface waters [6-8].

Due to these factors, there’s a growing interest in breast MRI approaches reducing GBCA administration in the screening context, yet providing visual characteristics of contrast-enhanced acquisitions by generating artificial contrast-enhanced images from non-enhanced acquisitions. Such methods were previously shown in breast studies [9-12], albeit deriving only one of the multiple time points considered in DCE breast MRI.

However, technically such methods might allow as well for generating images reflecting different time points of dynamic acquisitions by training distinct networks for each time points acquired over several minutes. This assumption is supported by several studies using different post-contrast acquisition intervals as training data. For example, in Chung et al. 90-second intervals [9], in Müller-Franzes et al. 60-second intervals [11], and in Kim et al. both 60 and 90 seconds [10] were used. These variations thus fuel and support the hypothesis of a principal possibility to predict post-contrast images across a series of time points in an individual patient. Furthermore, Zhang et al. [13] showed that besides synthesizing multiple different MRI sequences a feasibility to synthesize DCE in breast MRI is given. However, resulting in unsatisfying outputs as stated by the authors [13].

Our study therefore investigates neural networks’ ability to predict contrast enhancement for creating virtual dynamic contrast-enhanced (vDCE) breast MRI at various time points. It assesses two methods: using five separate networks for each time point (single-channel output - SCO-Net) and another using a single network with five time points as output channels (multi-channel output - MCO-Net), with the latter potentially benefiting by learning the dependencies of cross-time point latency information within a single training step. Both approaches were quantitatively assessed across the entire breast and in segmentations of findings, including delineation comparisons between second time point DCE and vDCE images, akin to Chung et al.’s work [9].

## 2 Materials and Methods

### 2.1 Dataset

This IRB approved retrospective study includes n=540 clinically indicated breast MRI examinations of female patients (mean age: 52±12 years) acquired between 2017 and June 2020 at University Hospital Erlangen, Germany. Acquisitions were conducted on a single 3 Tesla MRI scanner (MAGNETOM Skyra Fit, Siemens Healthineers, Erlangen, Germany) with a dedicated 18-Channel breast coil (Siemens Healthineers, Erlangen, Germany). The dataset was randomly split on patient level into training, validation and an independent test set with n=377, n=81, and n=82 examinations, respectively.

Each examination included pre-contrast T1w, T2-weighted (T2w), multi-b-value (b-values: 0, 750 and 1500 s/mm^2^) diffusion weighted imaging (DWI) and T1w subtraction series after administration of GBCA (T1w-sub) at five consecutive time points. The acquisition of T1w-sub began 20 seconds after GBCA injection with each scan lasting 60 seconds. Detailed acquisition parameters are provided in Table 1.

**Table 1.**
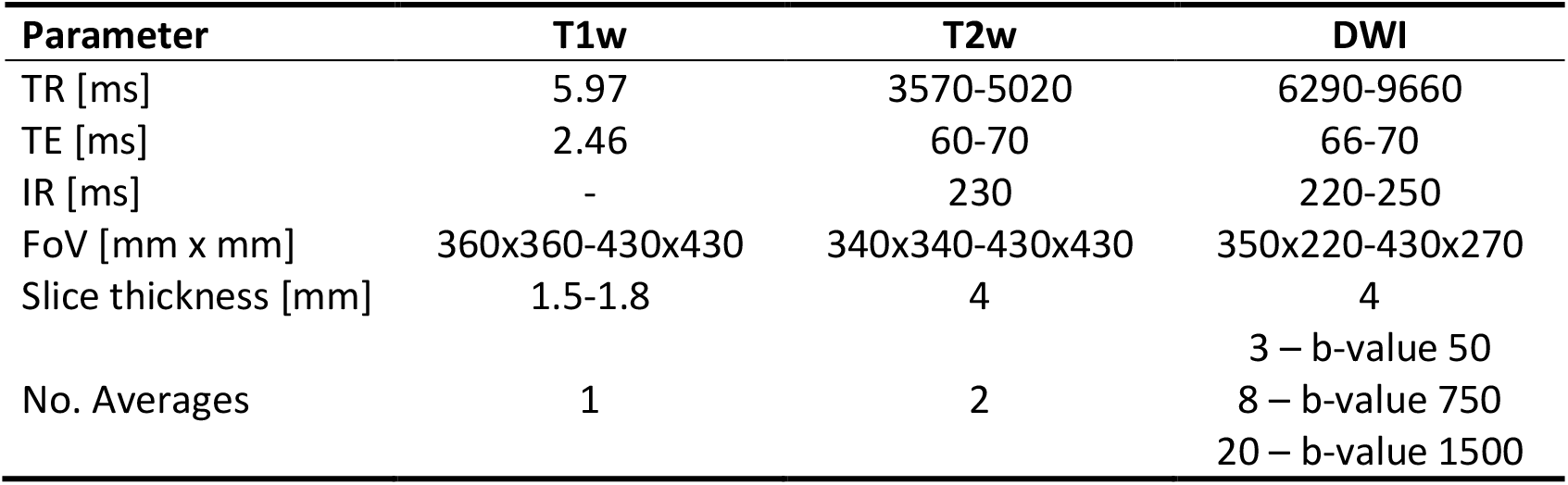
MRI Sequence Parameter. Repetition time (TR), echo time (TE), inversion recovery time (IR), field of view (FoV)

| Parameter | T1w | T2w | DWI |
| --- | --- | --- | --- |
| TR [ms] | 5.97 | 3570-5020 | 6290-9660 |
| TE [ms] | 2.46 | 60-70 | 66-70 |
| IR [ms] | - | 230 | 220-250 |
| FoV [mm x mm] | 360x360-430x430 | 340x340-430x430 | 350x220-430x270 |
| Slice thickness [mm] | 1.5-1.8 | 4 | 4 |
| No. Averages | 1 | 2 | 3 – b-value 50<br>8 – b-value 750<br>20 – b-value 1500 |

### 2.2 Data Preprocessing and Binary Masking

The DICOM files were transformed to NIfTI format using dcm2niix tool [14], followed by preprocessing using in-house Python (version 3.9.10) scripts utilizing SimpleITK framework (version 2.2.1) including resampling, intensity normalization, intensity clamping and rescaling.

All images’ field of view (FoV) was resampled to match the FoV of DWI acquisitions as well as common spatial dimensions with an in-plane matrix of 448×280 and n=96 slices. Z-score normalization was applied individually on T1w, T2w and different DWI acquisitions. To maintain the intensity uptake over time in T1w-sub normalization was applied to the entire series. Intensities were clamped at boundaries of −1 and 15, to minimize outlier impact. Finally, T1w, T2w and the DWI acquisitions were scaled to a [0, 1] domain, while T1w-sub volumes were scaled to a [−1,1] domain.

Binary breast volume masks were calculated from T1w data using an in-house developed algorithm based on mean thresholding of multiple maximum intensity projections (MIP) in slice (z-) direction. Each *slice* of T1w data was processed into individual MIPs of adjacent slices (*n*_*slices*_) to account for varying anatomical shapes, as defined in Equation (1). MIPs were then binarized using mean thresholding followed by binary dilation with a 5-pixel diameter disk-shaped kernel and a binary closing operation to ensure homogeneous masks. The resulting masks were stored in NIfTI format with the same spacing, direction and origin of the T1w data. This algorithm was also previously described in Liebert et al. [15].

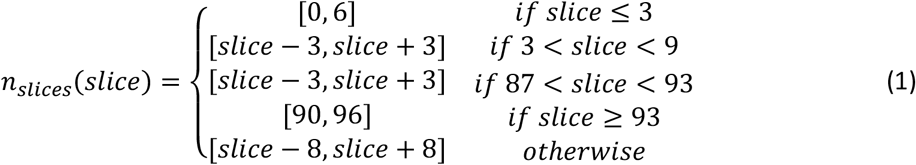

### 2.3 Neural Network Architecture and Training

Two 2D U-net architectures, were implemented, both consisting of three encoder and three decoder stages with a bottleneck layer in-between. Detailed information of the composition of each layer is presented in Figure 1.

**Fig. 1.**
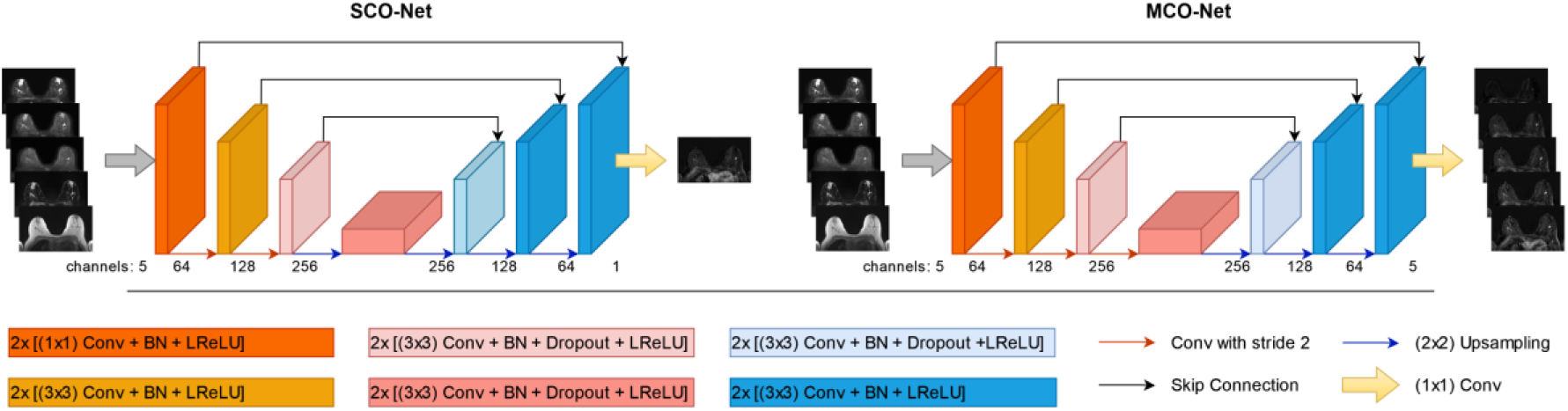
Schematic overview of 2D-Unet Architectures. Left: Architecture for SCO-Net. Right: Architecture for MCO-Net. Both networks use 5 images as input channels. In SCO-Net, five individual networks were trained, each using a different time point of the DCE series as ground-truth. The MCO-Net uses each time point of the DCE series as a channel of the ground truth. Encoder stages consisted of two blocks of either a 1×1 or 3×3 convolution (Conv) layers followed by a batch normalization (BN) layer and a leaky rectified linear unit (LReLU) layer. Each decoder stage incorporates a concatenation with features of the encoder to represent the skip connections. The decoder stages on the 2nd and 1st level consisted of two 3×3 Conv layers followed by a BN layer and a LReLU layer. The deepest encoder and decoder stage and the bottleneck contain an additional dropout layer with a probability of 0.5. For encoder down-sampling, 2×2 Conv with a stride of 2 were used. Decoder up-sampling was performed using 2×2 transposed Conv with a stride of 2. After the final decoder stage, a 1×1 Conv was performed followed by a *tanh* operation in order to map the predictions to the expected output channels and [−1, 1] domain.

In SCO-Net, five individual networks were trained each predicting a single time point of the DCE series. MCO-Net utilized a five-channel output representing the whole DCE time series. The networks were trained using native T1w, T2w and the multi-b-value DWI series as inputs and T1w-sub series as targets. Inspired by Chen et al.[16] the loss function was a combination of structural similarity index metric (SSIM)[17] and L1-norm as shown in the Equation (2) below.

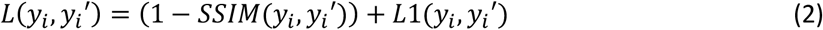

Training utilized all slices of the datasets with ADAM optimizer and a batch size of 30 random slices. A dedicated workstation (Linux Ubuntu 20.04, AMD Ryzen Threadripper PRO 3945WX 3.4Ghz, 64GB RAM) with one Nvidia Quadro RTX 6000 GPU-card with 24GB RAM was used. The networks were trained for 35 epochs without early stopping. Experiments were implemented using PyTorch (version 1.13.1), PyTorch-Lightning (version 1.8.6) and MONAI (version 0.8.0) frameworks.

### 2.4 Performance Evaluation

The vDCE series were quantitatively evaluated on the holdout test set using the following metrics also used in previous literature: SSIM, peak signal-to-noise-ratio (PSNR), normalized root mean square error (NRMSE) and median symmetric accuracy (MEDSYMAC) which addresses robust analysis of symmetric prediction errors [10,12,18,19]. These metrics were calculated for both the whole image volume and separately for bounding boxes placed around segmented target findings as detailed below. Additionally, high frequency error norm (HFEN) was calculated only for whole images volume, as target finding sizes were too small in some cases for the calculation. Differences in mean values per time point and subject between the two network architecture approaches were evaluated using a two-way Repeated Measurements Anova, with the pingouin Python framework (version 0.5.3).

To assess the proposed techniques’ effectiveness in depicting contrast uptake, segmentations were conducted on n=48 subjects of the test set with findings that stood out against background tissue in the DCE acquisitions including 8 BI-RADS<=2 cases (ranging from 5.0-31.4mm) and 40 BI-RADS>2 cases (ranging from 4.7-85.1mm). Per patient, the largest appearing finding was volumetrically segmented in DCE and both vDCE methods at the second time point using 3D Slicer Software (version:4.11) [20] carried out by a medical student (>2 years’ experience) under a single radiologist’s (>10 years’ experience) guidance. Segmentations were compared using following metrics: DICE score, Intersection over union (IoU), Hausdorff Distance (HD), and segmentation volume (segV). Additionally, the contrast-to-noise ratio (CNR) was evaluated using a bounding box with a one-pixel offset around segmentations. Statistical analysis was performed using a one-way Repeated Measures ANOVA with scipy (version 1.9.1) considering a p-value<0.05 as significant.

## 3 Results

### 3.1 Binary Masking

Figure 2 displays the results of breast volume masking on corresponding T1w slices for a representative case. The masks successfully include the whole breast tissue while the air around the patient is excluded as well as parts of the thorax, and most of the lung and heart tissue.

**Fig. 2.**
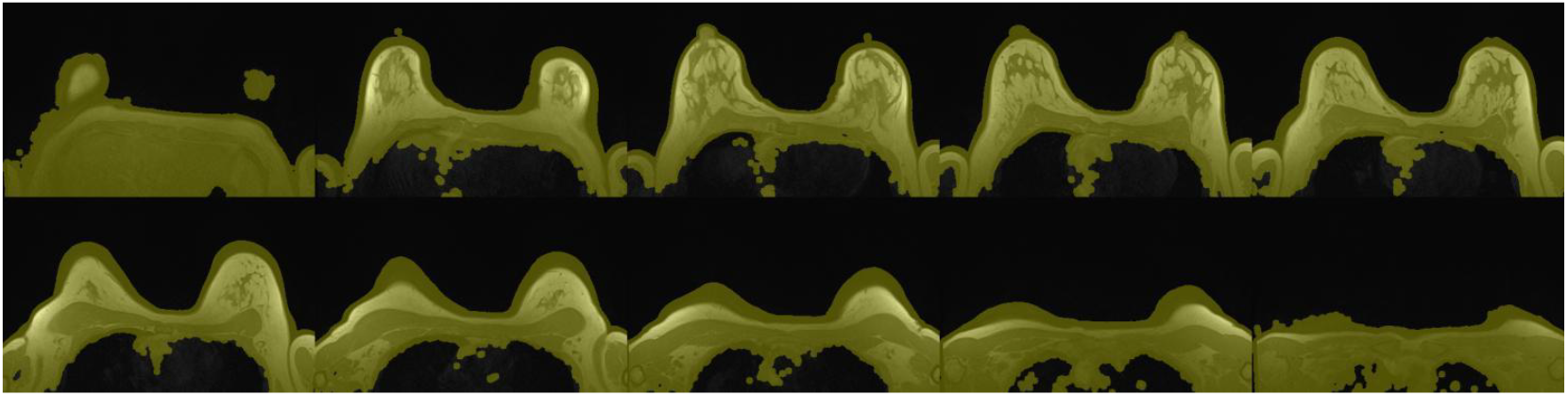
Exemplary case of the breast volume masking for ten different slice positions is displayed. T1w slices and the corresponding binary mask overlay (yellow) are shown. The mask correctly excludes air around the body and large parts of the lungs. Whereas the breast tissue is fully included with an additional offset around the outer breast contours to ensure no important information is counted as background

### 3.2 Neural Network Performance

Generated vDCE images were evaluated on similarity and error metrics for both SCO-Net and MCO-Net on the holdout test set. Mean values across all five time points are presented in Table 2. SCO-Net reached marginally higher values for SSIM and marginally lower values for PSNR and HMI in the image volume and mean NRMSE and HFEN values were lower for MCO-Net and MEDSYMAC higher. In the segmentations, MCO-Net showed higher similarity and lower error metrics compared to SCO-Net except MEDSYMAC.

**Table 2.** Mean (±Std) similarity and error metrics of vDCE in image volume and segmentations of SCO-Net and MCO-Net.

|  | Image Volume |  | Segmentation |  |
| --- | --- | --- | --- | --- |
|  | SCO-Net | MCO-Net | SCO-Net | MCO-Net |
| SSIM (↑) | 0.864±0.025 | 0.863±0.026 | 0.591±0.133 | 0.596±0.131 |
| PSNR [dB] (↑) | 24.288±2.379 | 24.424±2.416 | 22.065±3.448 | 22.204±3.395 |
| HMI (↑) | 0.694±0.066 | 0.701±0.066 | 1.476±0.741 | 1.481±0.737 |
| NRMSE (↓) | 0.089±0.015 | 0.087±0.016 | 0.174±0.049 | 0.173±0.049 |
| MEDSYMACE (↓) | 0.020±0.011 | 0.022±0.012 | 0.106±0.032 | 0.105±0.032 |
| HFEN (↓) | 0.796±0.079 | 0.780±0.073 | - | - |
| SSIM – structural similarity index measure, PSNR – peak signal to noise ratio, HMI – histogram mutual information, NRMSE – normalized root mean square error, MEDSYMACE – median symmetric accuracy, HFEN – high frequency error norm. |  |  |  |  |

Furthermore, similarity and error metric values separated per time point for each of the above-described setups are presented in Figure 3. It can be noted that the later time points result in significantly higher SSIM and HMI values and lower PSNR. These trends are visible both in the image volume and segmentations. Error metrics show significantly lower values for later timepoints.

**Fig. 3.**
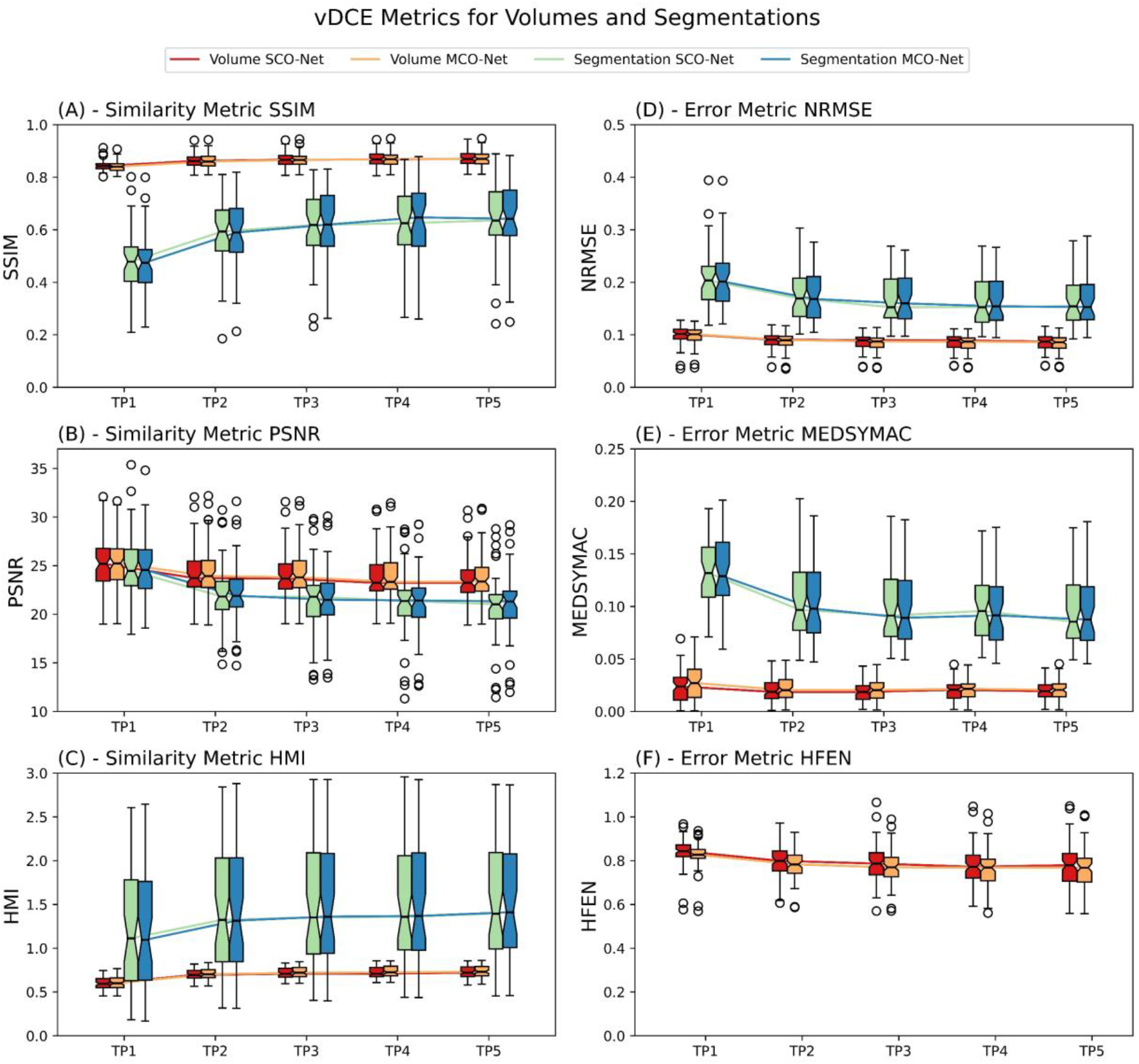
Similarity metrics for vDCE SCO-Net and MCO-Net in image volume as well as segmented findings are presented. SSIM values show an increase and PSNR values a decrease for later time points both for the volume and segmentations.

The metrics in Table 2 and Figure 3 indicate a generally worse performance inside segmentations compared to image volume. SSIM indicated no statistically significant (p>0.05) differences between SCO-Net and MCO-Net in image volume. However, all other metrics showed significant effects (p<0.05) between the two approaches. Significant differences (p<0.05) were found for SSIM, PSNR, HMI and MEDYSMAC across different time points and approaches in image volume. NRMSE and HFEN showed non-significant differences (p>0.05) across different time points.

In Figure 4 five consecutive time points of three representative cases of both vDCE approaches and of the original DCE series are shown. In both vDCE approaches the localization of the lesion is well correlating with the DCE images as well as the signal intensity uptake over time.

**Fig. 4.**
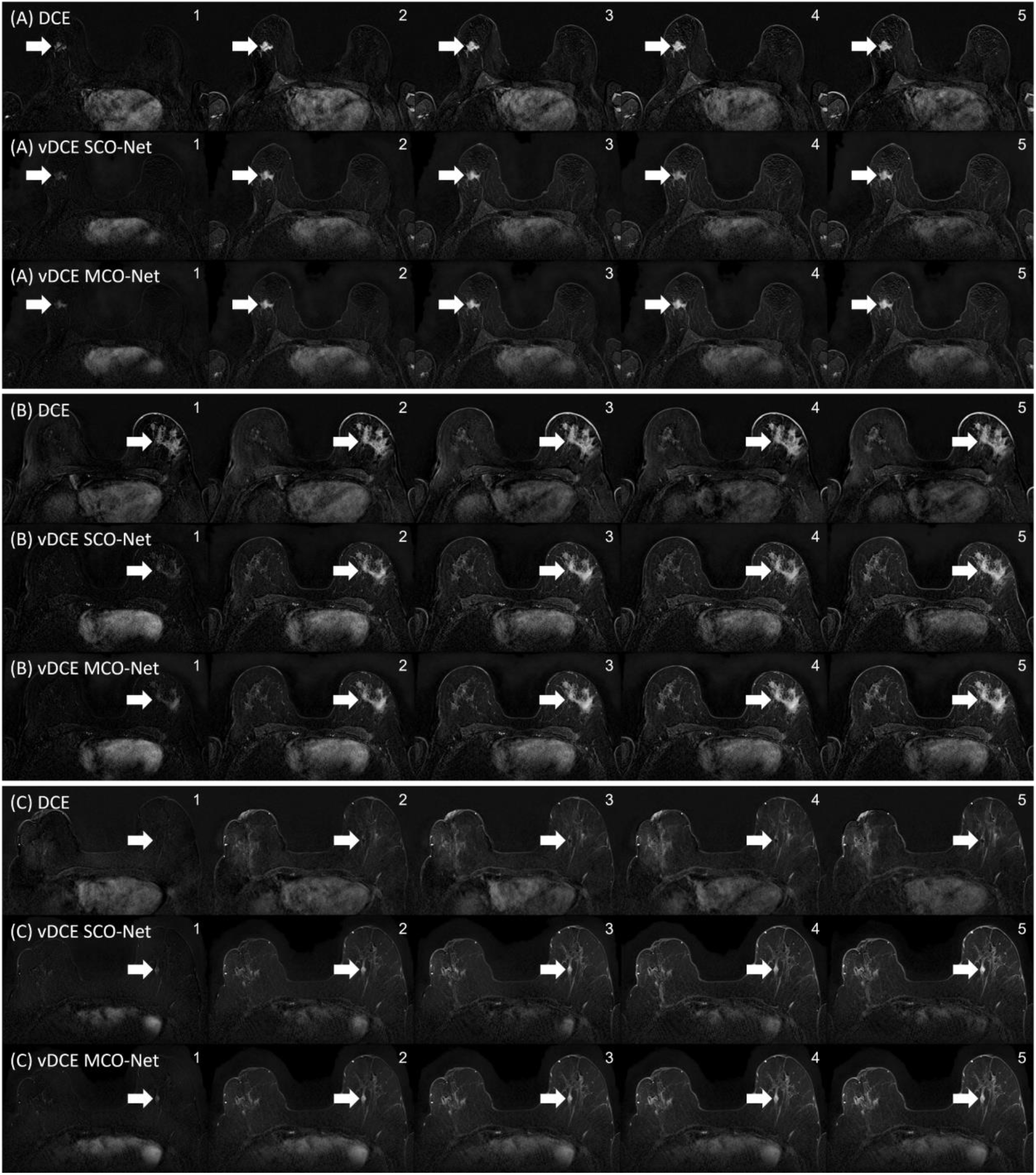
(A) - Exemplary case of a 60- to 70-year-old patient with a histopathologically confirmed malignant lesion in the right breast. (B) - Exemplary case of a 60- to 70-year-old patient with a histopathologically confirmed non-mass enhancement in the left breast. (C) Exemplary case of a 50- to 60-year-old patient having a cyst in the left breast. Respectively for each case: Top: DCE series, Middle: vDCE SCO-Net, Bottom: vDCE MCO-Net. In each series the depiction of the lesions is visible with an increasing signal intensity over time. In (A) stronger lesion enhancement can be observed in the DCE image when compared to the vDCE approach. A weaker enhancement appears for the 4th time point of the vDCE SCO-Net when compared to both the vDCE MCO-Net and DCE series. (C) shows a false-positive enhancement in both vDCE approaches as cyst do not enhance in DCE series.

### 3.3 Evaluation of Ability to Depict Target Findings

Figure 5 displays three lesions with its corresponding segmentations performed on DCE as well as vDCE SCO-Net and MCO-Net. The lesion shapes and location appear similarly. However, this also shows how lesion depiction appears differently for the different approaches.

**Fig. 5.**
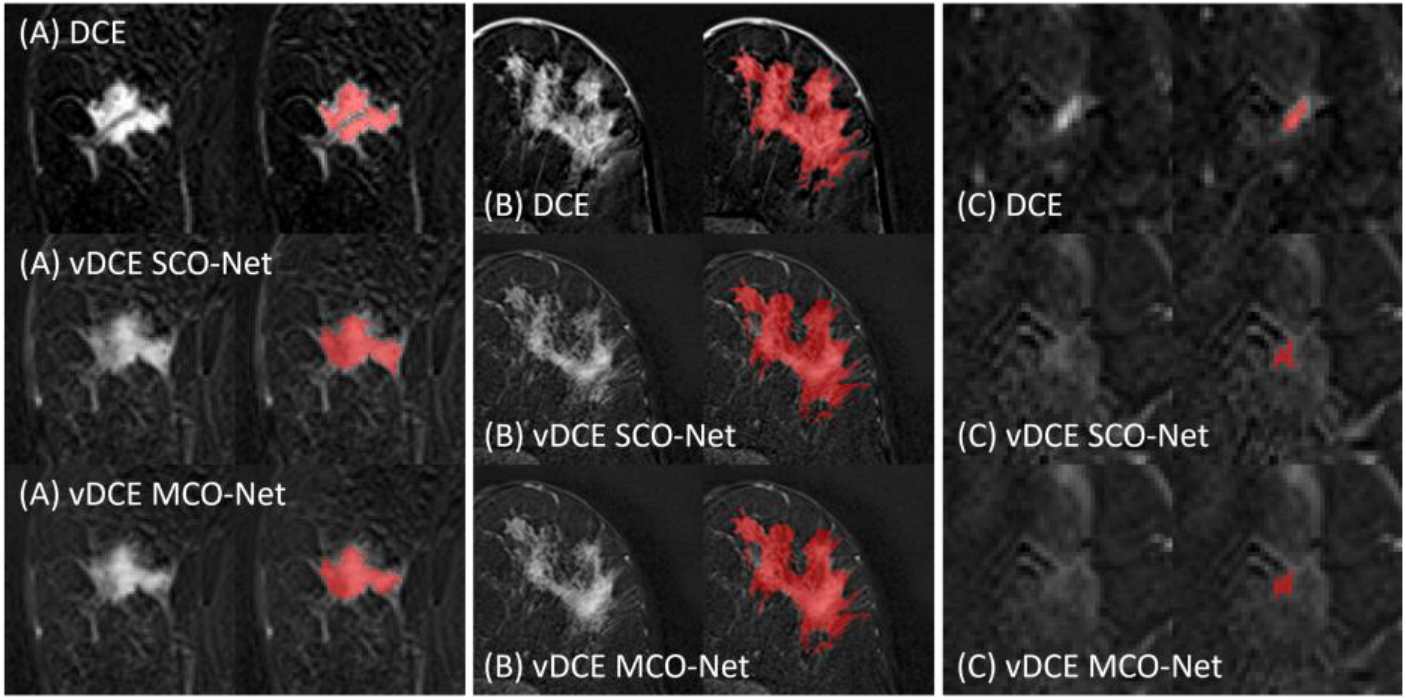
Segmentation differences of lesions. Top: DCE image, Middle: vDCE SCO-Net, Bottom: vDCE MCO-Net. (A) represents a malignant lesion, (B) a non-mass enhancement and (C) a malignant lesion.

Table 3 shows the quantitative evaluation of segmentation correctness using SCO-Net and MCO-Net for vDCE generation. MCO-Net shows marginally higher mean DICE scores and IOU values and a lower HD. For the original DCE series a CNR=2.483±1.350 could be reached. This wasn’t significantly different from the CNR values reached by both of the vDCE approaches. No significant changes (p>0.05) were observed between SCO-Net and MCO-Net for any of the investigated segmentation metrics. Figure 6 shows the Bland-Altman plot for the segmented volume segV.

**Table 3.**
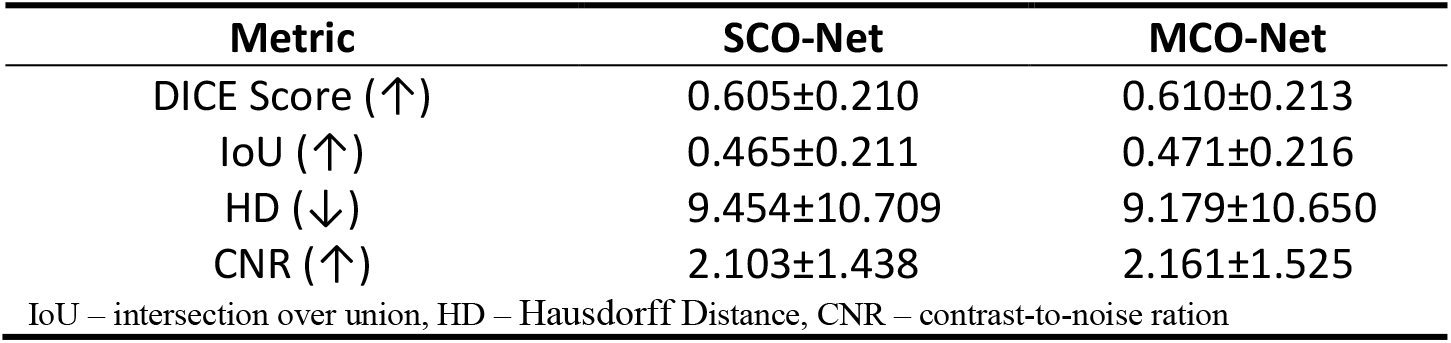
Quantitative Mean (±Std) Segmentation Metrics.

**Fig. 6.**
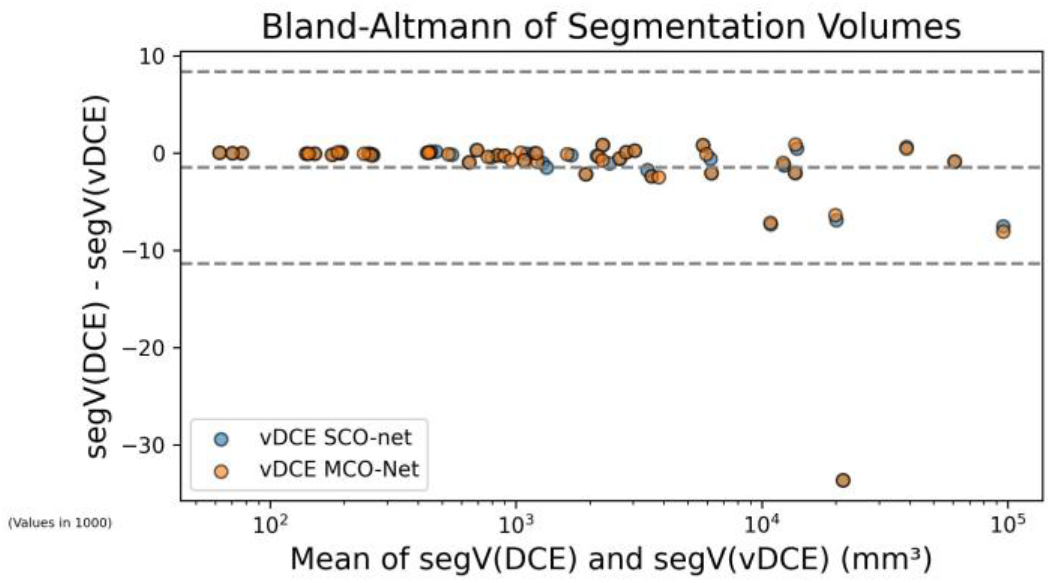
Bland-Altmann plot compares findings’ segV in the 2^nd^ time point of the DCE and vDCE series. An increase in variation in larger segmentations for both vDCE approaches, not meeting the DCE segV observed similarly for SCO-Net and MCO-Net. Overall, there is a systematic bias of −1.49 for vDCE approaches indicating underestimation of finding sizes, although this bias diminishes for smaller segmentations. Outlier segmentation case is shown in Figure 5 (C).

## 4 Discussion

This study evaluates the technical feasibility of generating a vDCE series for breast MRI comprising individual images for each time point post GBCA injection using two neural network approaches. One approach was trained on tissue characteristics at a specific time point, blinded to information of other time points (SCO-Net), while the other was trained to consider the entire dynamic cycle (MCO-Net), potentially benefiting from exploring cross-timepoint latency information. Both approaches depicted image characteristics similar to DCE images. MCO-Net showed a higher performance with significant differences across metrics except the SSIM compared to SCO-Net, in both image volume and segmented benign and malignant findings.

For SCO-Net, lowest SSIM scores and highest error metrics were observed for the 1^st^ time point of the DCE series, gradually increasing towards the later time points. The SCO-Net trained for the 2^nd^ DCE series time point, which correlates to an acquisition approx. 80 seconds after GBCA administration, reached a mean SSIM of 0.864±0.025 aligning with prior studies by Chung et al. [9] and Kim et al. [10], both trained networks on contrast-enhanced breast MRI data acquired approx. during this timeframe. Mean SSIM scores across all time points (0.84-0.87) align with previously noted literature values of 0.76-0.91 [9,10,12,13,18], while PSNR scores 23.64-25.37dB fall to lower end of previously noted literature ranges of 23.18-54.8dB [10,12,13,18]).

Despite minimal metric differences, a more homogenous tissue appearance over time points was observed when using MCO-Net (see Figure 4). The MCO-Net approach aimed to incorporate contextual information of time dependency of contrast-enhancement during training. It utilizes the interdependent information of various post-contrast images and their time dependency of different tissue compartments in the breast. The MCO-Net achieved higher PSNR and lower error metrics compared to SCO-Net and non-significant differences in SSIM. The improvement in SSIM and error metrics over time might correlate with higher signal in later time points. Despite this higher signal, the increased noise level in tissue, decreased PSNR due to its sensitivity to noise.

Compared to Chung et al.’s study [9], our segmentations yielded a lower DICE coefficient between DCE and vDCE approaches. This might be attributed to including more challenging cases with all findings independent of malignancy or mass/non-mass enhancement type. Incorporating segmentations into the loss function, as demonstrated by Chen et al. [16], may enhance performance on finding-delineation and signal intensity representation. However, in this study, neither network approach significantly outperformed the other in regards of comparing manual segmentations.

Utilizing 2D Networks increased dataset size and reduced resources needed for training networks with high resolution images. Still, the images showed high inter-slice homogeneity in z-direction. Exploring 3D architectures for vDCE prediction might further improve capabilities of such technique by incorporating information of adjacent slices.

Our study has limitations, including the absence of a qualitative reader study to assess clinical applicability, evaluations on enhancement kinetics over time and a potential segmentation bias. Further clinical evaluations should be pursued in future research with larger cohorts. Additionally, the study’s reliance on data from a single MRI scanner model limits the generalizability of our findings, emphasizing the need for research incorporating diverse equipment. Future work should also investigate which MRI sequences are required for vDCE generation, in analogue to investigation of Liebert et al. [21] to further explore dependency on acquisition techniques and generalizability.

## 5 Conclusion

In conclusion, providing a neural network information on tissue enhancement kinetics during training in form of additional input channels, like in MCO-Net approach, significantly improved metrics for generating a time-dependent series of vDCE breast MRI images from unenhanced acquisitions. Further research in this area might be justified based on these findings and should incorporate individual assessment of lesion specific enhancement characteristics as clinically relevant targets.

## Data Availability

All data used and produced in the present study are not publicly available due to preservation of personal privacy under the european GDPR. Institution handling this data is the Radiological Department of University Hospital Erlangen.

## Acknowledgments

This project is funded by the Bavarian State Ministery of Science and Arts in the framework of the bidt Graduate Center for Postdocs. The work was performed in (partial) fulfillment of the requirements for obtaining the degree "Dr. rer. biol. hum.” at the Friedrich-Alexander-Universität Erlangen-Nürnberg (FAU).

